# Severe Mitral Regurgitation in Paradoxical Low-Flow Low-Gradient Severe Aortic Stenosis

**DOI:** 10.1101/2024.09.19.24314018

**Authors:** Shani Dahan, Jacob Dal-Bianco, Ygal Plakht, Mayooran Namasivayam, Romain Capoulade, Xin Zeng, Jonathan J Passeri, Evin Yucel, Michael H Picard, Robert A Levine, Judy Hung

**Affiliations:** Division of Cardiology, Department of Medicine, Massachusetts General Hospital, Harvard Medical School, Boston, Massachusetts; Department of Nursing, Faculty of Health Sciences, Ben-Gurion University of the Negev, Beer-Sheva, Israel; Department of Cardiology, St. Vincent’s Hospital, Sydney, Australia; Quebec Heart and Lung Institute, Laval University, Quebec City, Quebec, Canada

**Keywords:** Aortic Stenosis, Mitral Regurgitation, Aortic Valve Replacement, Valve Disease, Transvalvular Flow Rate

## Abstract

**Background:** Patients with paradoxical low-flow, low-gradient severe aortic stenosis (LFLGAS) exhibit low transvalvular flow rate (Q), while maintaining preserved left ventricular ejection fraction (LVEF). Concomitant severe mitral regurgitation (MR) contributes to the low flow state, adding complexity to diagnosis and management. This study aimed to examine the impact of severe MR on outcomes in paradoxical LFLGAS.

**Methods:** Data from an institutional echo database identified 1,189 patients with adjudicated severe aortic stenosis (AVA≤1.0 cm^2^), low transaortic gradients (mean gradient<40 mmHg), preserved LVEF (≥50%), and low flow rate (Q≤210 ml/sec), to confirm paradoxical LFLGAS. Subgroups were based on MR severity (severe and non-severe). Clinical outcomes included all-cause mortality, aortic valve replacement (AVR), heart failure hospitalizations, and a composite outcome.

**Results:** In the severe MR group (n=80), patients had lower flow rates, increased LV dimensions and a more eccentric hypertrophy pattern compared to non-severe MR (n=1,109). Over a median 5-year follow-up, severe MR correlated with higher all-cause mortality (p=0.02) and AVR rates (p=0.012). After adjustment, severe MR was independently associated with increased all-cause mortality risk (HR=1.43, p=0.011) and composite outcome (HR=1.64, p<0.001). AVR significantly reduced mortality at every MR degree, with the most substantial impact in severe MR (HR=0.18, p<0.001). Propensity-adjusted models demonstrated a stronger AVR impact with increasing MR degree (p-for-interaction=0.044).

**Conclusions:** Severe MR in paradoxical LFLGAS is associated with adverse outcomes and distinctive LV remodeling. Aortic valve replacement improves survival across all MR grades, with greater impact in severe MR.

## Introduction

Patients with severe aortic stenosis may present with low transaortic flow rate and gradients due to reduced stroke volume, despite preserved left ventricular ejection fraction (LVEF). This condition, referred to as paradoxical low-flow, low-gradient severe aortic stenosis (LFLGAS) (1), represents a diagnostic and therapeutic challenge in daily clinical decision making. The challenges arise from uncertainties regarding the true severity of the stenosis, the underlying pathophysiological processes that define this condition and the preferred treatment strategy.

Additionally, these patients are often misclassified as having moderate aortic stenosis, leading to less frequent or delayed surgical referrals, and ultimately poorer clinical outcomes (2–5).

The development of the low flow state in LFLGAS is generally attributed to a restrictive physiology characterized by concentric left ventricular (LV) hypertrophy, resulting in a smaller LV cavity size, impaired LV filling and consequent diminished flow across the aortic valve (6,7). In addition, intrinsic impairment of myocardial contractility, mostly decreased longitudinal function, has been shown to contribute to this hemodynamic profile (8).

Mitral regurgitation (MR), which commonly occurs with severe aortic stenosis, impairs forward flow by diverting a substantial fraction of the total LV stroke volume backwards through the mitral valve into the left atrium, instead of forward across the aortic valve. Significant MR has been previously shown to be an independent factor contributing to the low flow state in LFLGAS (9,10), and it can affect the diagnostic accuracy of flow-dependent aortic valve area (AVA) measurements (11,12). While paradoxical LFLGAS and MR frequently coexist in clinical settings, the optimal treatment strategy remains unclear, given that the effects of severe MR on clinical outcomes are currently unknown.

The aim of our study was to investigate the echocardiographic remodeling patterns and the clinical outcomes in patients presenting with paradoxical LFLGAS and simultaneous severe MR, as well as the effect of aortic valve replacement (AVR) on survival in this important patient population.

## Methods

### Study cohort

We included patients who underwent echocardiography at our institution between the years 2001 and 2022 and met the criteria for paradoxical low-flow, low-gradient severe aortic stenosis. This was determined based on a calculated aortic valve area (AVA≤1.0 cm^2^), transaortic gradients (mean gradient ≤40 mmHg), LVEF≥50% and flow state. To define flow state, we calculated transvalvular flow rate (Q) for each patient, utilizing a mathematical equivalent method derived from mean aortic valve gradient and peak aortic valve velocity, as previously described (13). We defined low flow state as Q≤210 ml/sec (6,13–15) to confirm the diagnosis of paradoxical LFLGAS. Patients with prosthetic aortic valve, supravalvular or sub-valvular aortic stenosis, were excluded.

### Echocardiographic data

The echocardiographic data for each patient was sourced from their initial available echocardiogram during the study period. Quantitative data, including aortic valve area calculation, were derived from the values documented in the official clinical read. All readers possessed a level III certification in echocardiography. MR was graded as mild, moderate or severe using a multiparametric integrative approach, as recommended in the current American Society of Echocardiography guidelines (16). LV mass was calculated using the linear method (17), and relative wall thickness was calculated using the recommended formula: (2*posterior wall thickness)/(LV internal diameter at end diastole) (17).

### Clinical data

The clinical characteristics, including comorbidities, were determined from review of the institution’s electronic health records. Aortic and mitral valve procedures, both open surgery and transcatheter approaches, were identified using the Current Procedural Terminology coding system. Death from any cause was determined at 5 years from the time of the index echocardiogram in each patient. Mortality data, including dates of death, were obtained from the electronic health record, which integrates social security and clinical death records.

### Follow-up and study outcomes

The follow-up lasted up to 5 years from the time of the index echocardiogram. The primary outcome was all-cause mortality. The secondary outcomes included AVRs: surgical or transcatheter aortic valve replacement (SAVR/TAVR) and heart failure (HF) hospitalizations. A composite cardiovascular outcome was also defined, encompassing all three. Individuals were either excluded or censored from the survival analysis if they underwent mitral valve intervention before or after the AVR, respectively.

### Statistics

Categorical variables were expressed as a number (percentage) and continuous variables as mean ± standard deviation (SD) / median with interquartile range (IQR). Comparisons between the study groups in univariate level were performed using Chi-squared test for categorial variables and Student’s *t*-test for count data. Probabilities of the study outcomes were investigated using survival approach (Kaplan-Meier method) and compared with Log-Rank test. The associations between MR and the risk for the investigated outcomes, in univariate and multivariate levels, were assessed using Cox Proportional Hazard Regression models. The multivariable models included the baseline characteristics clinically and statistically related to the outcomes. In addition, we included the results of a propensity score in the models. The propensity score was built based on the following parameters clinically associated with high likelihood for AVR: age, sex, AVA, body surface area, diabetes mellitus, atrial fibrillation, chronic kidney disease, history of heart failure, myocardial infarction, and stroke. Interaction analyses were conducted to assess the specific correlation of AVR with all-cause mortality within different subgroups based on MR severity. For each test, a p-value (two-sided) <0.05 was considered significant. Analyses were performed using Statistical Package for the Social Sciences (SPSS), version 29 (IBM Corporation, Armonk, NY, USA).

### Ethics approval

The study received ethical approval from the Massachusetts General Hospital/Partners Institutional Review Board. Informed consent was not required.

## Results

### Study cohort

The overall study cohort included 1,189 patients, of which 388 (32.6%) were male, with a mean age of 83.0±8.6 years. The mean Q was 173.7 ± 25.6 ml/s (confirming a low flow state) and the mean LVEF was 64.1±7.9% for the entire cohort. MR grades on index echocardiography were mild or less (n=480), moderate (n=629) and severe (n=80). The study population was divided into two groups according to MR severity on index echocardiography: patients with severe MR (n=80 patients [6.7%]), and patients with non-severe MR (n=1,109 [93.3%]) (Figure 1).

**Figure 1:**
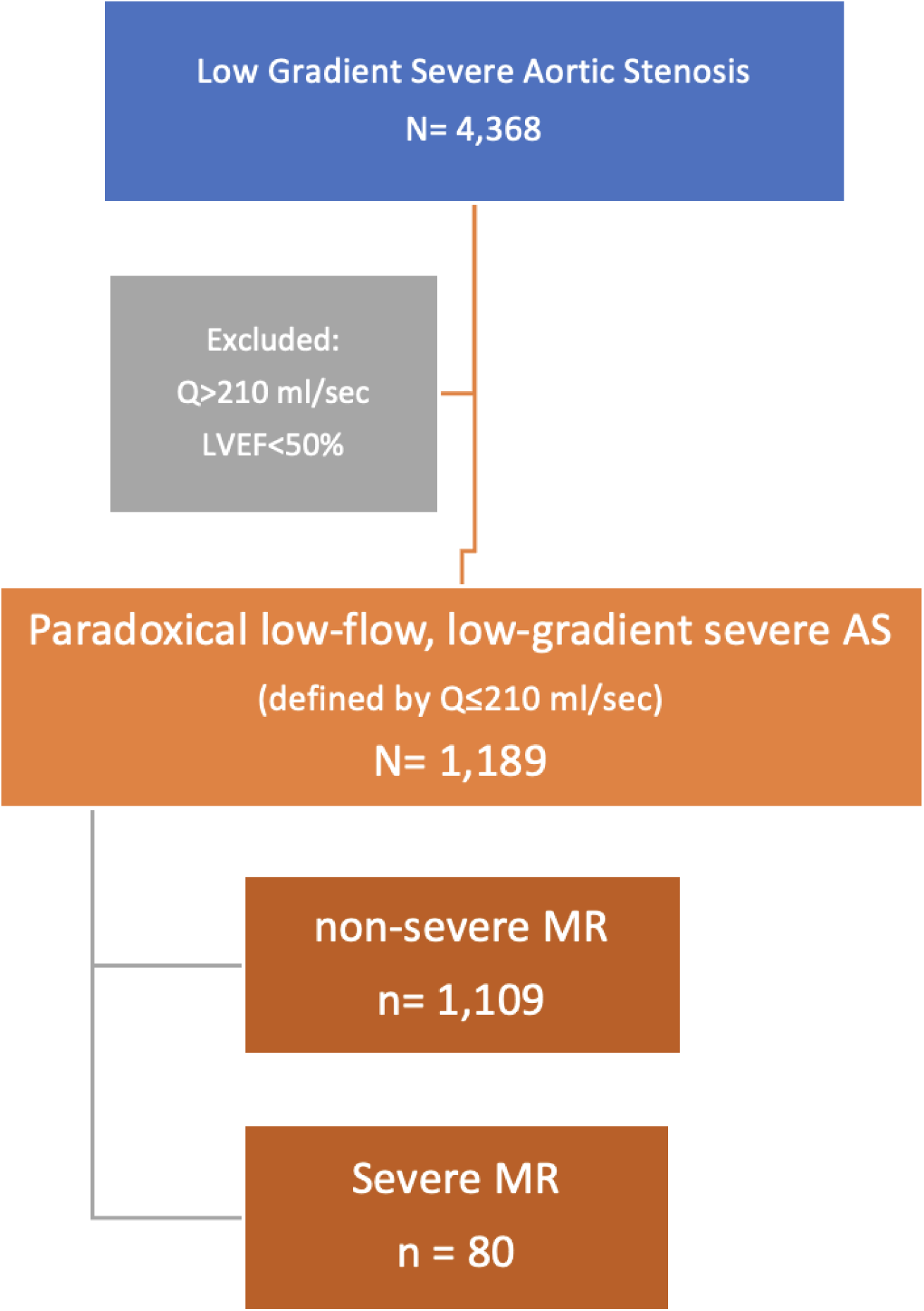
Flow Chart of Patient Selection: This flow chart outlines the selection process for patients with paradoxical low-flow, low-gradient severe aortic stenosis (LFLGAS). Patients were identified from an institutional echocardiography database using the following criteria: AVA ≤1.0 cm², mean gradient ≤40 mmHg, LVEF ≥50%, and low transvalvular flow rate (Q ≤210 ml/sec). The study population was then classified into two groups, based on mitral regurgitation (MR) severity: severe MR (n=80) and non-severe MR (n=1,109). *AVA = aortic valve area; LVEF = left ventricular ejection fraction; MR = mitral regurgitation; Q = transvalvular flow rate*.

Table 1 displays the clinical characteristics of the two study groups. Patients with severe MR were older (p=0.039) compared to those with non-severe MR, but there was no difference in comorbidities between the two groups. Echocardiography data revealed that patients with severe MR had lower flow rates compared to patients with non-severe MR (p<0.001). Additionally, severe MR was associated with larger left ventricular end-diastolic (p<0.001) and end-systolic (p=0.002) dimensions, higher LV mass index (p<0.001) and lower relative wall thickness (p=0.049), consistent with a more eccentric remodeling pattern (Table 1).

**Table 1:**
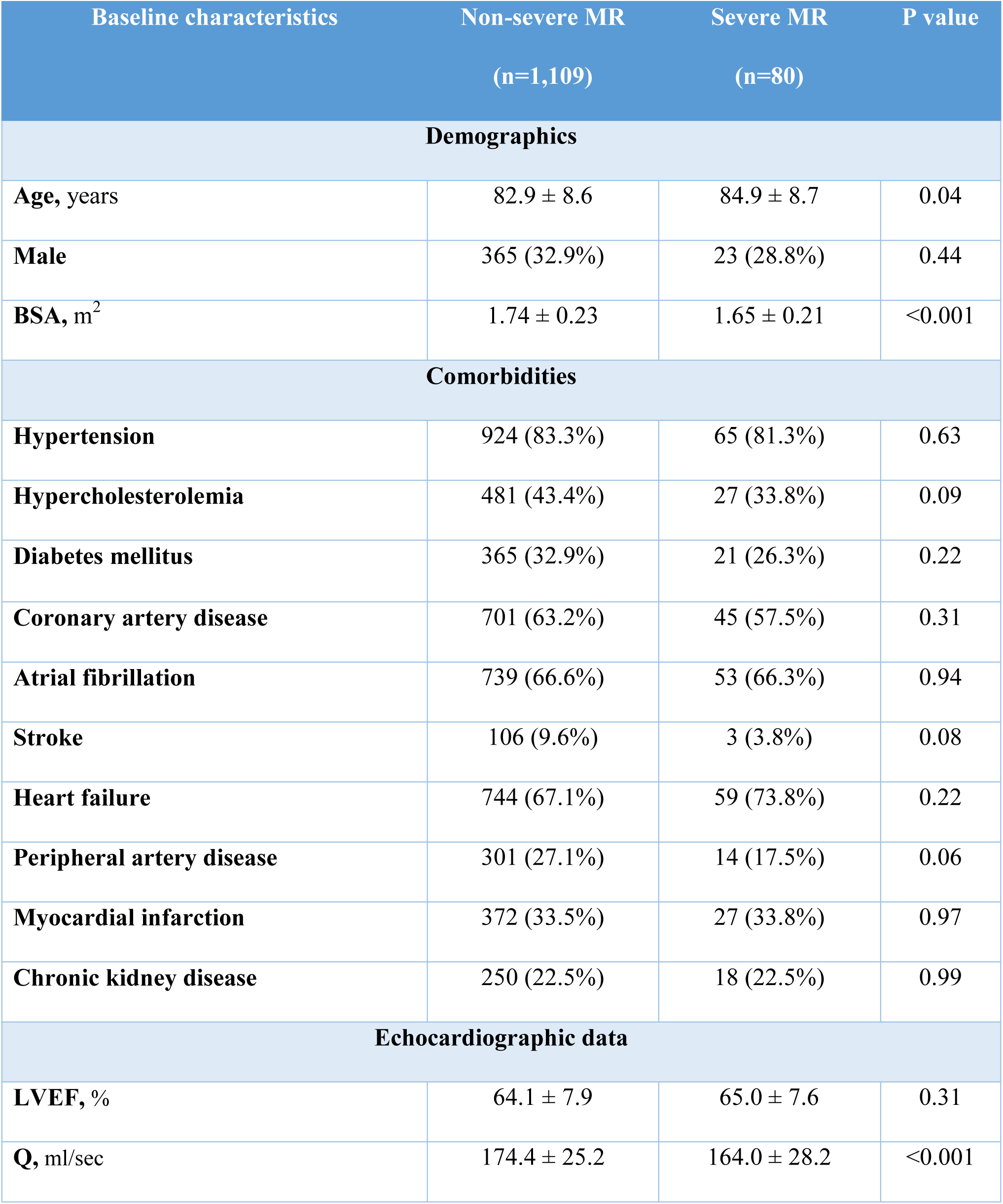
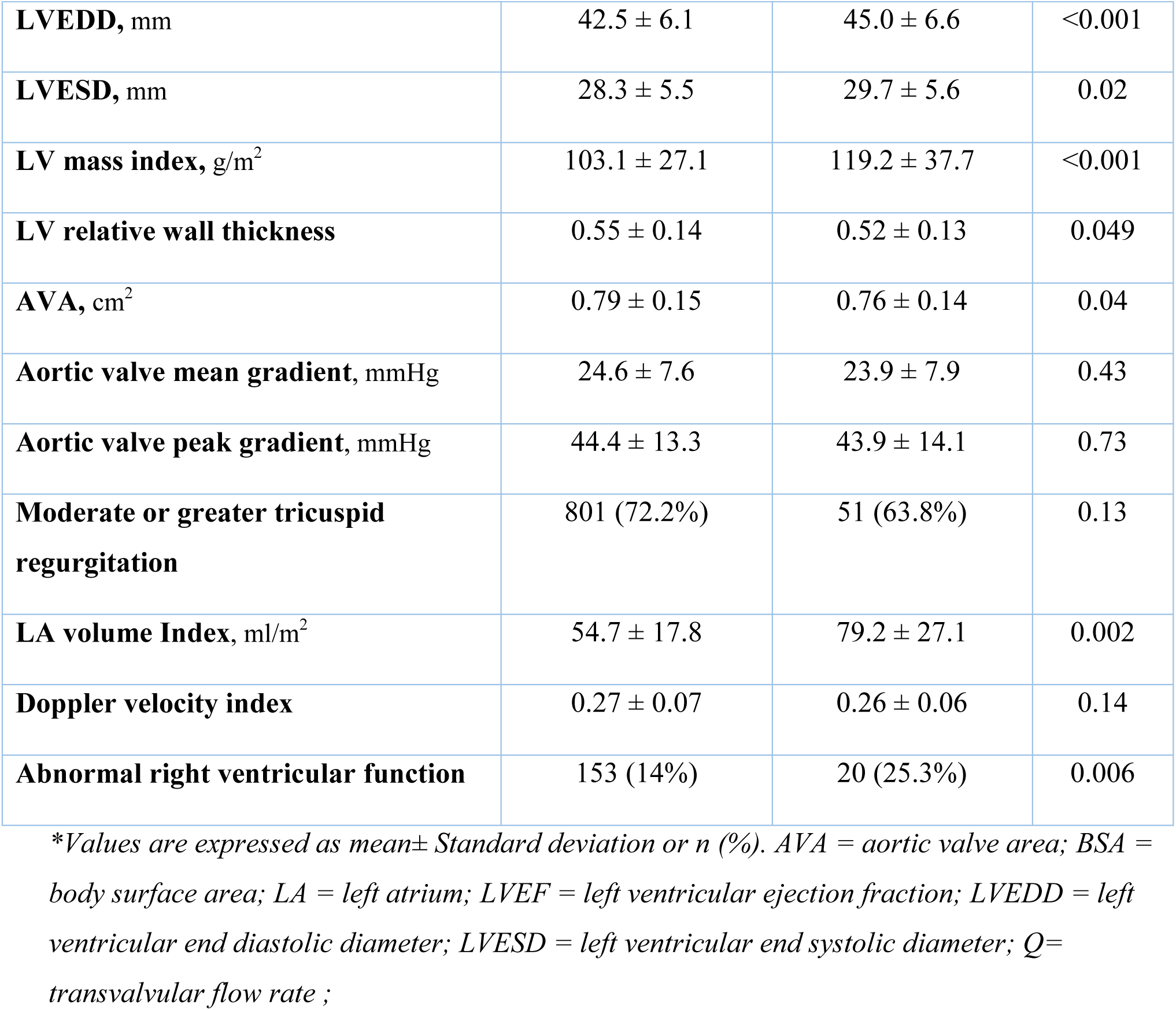
Baseline Characteristics.

### Survival analysis

During a follow-up period of up to 5 years, with a median duration of 0.93 years (equivalent to 246 days, IQR: 39-770 days), 947 events were detected, accounting for 79.6% of the entire cohort. This included 721 all-cause deaths (60.6%, cumulative incidence 0.71), 118 AVRs (9.9%, cumulative incidence 0.17), and 434 HF hospitalizations (36.5%, cumulative incidence 0.56). Mortality rates (and cumulative mortalities) according to the severity of MR were as follows: mild MR – 63.1% (0.77), moderate MR – 61.0% (0.72), and severe MR – 70% (0.84). A significant difference in cumulative mortality was found between severe MR and moderate MR (p=0.005) and between severe MR and mild MR (p=0.034) (Supplemental Figure 1).

Patients with severe MR had increased cumulative incidence of all-cause mortality (0.83 vs 0.70, p=0.002), need for AVR (0.32 vs 0.17, p=0.012), and a composite outcome of all-cause mortality, AVR and HF hospitalizations (0.98 vs 0.89, p<0.001), compared to those with non-severe MR (Table 2, Figure 2, Figure 3). However, there was no significant difference in the incidence of HF hospitalizations between the two groups (0.54 vs 0.56, p=0.353) (Figure 3b).

**Figure 2:**
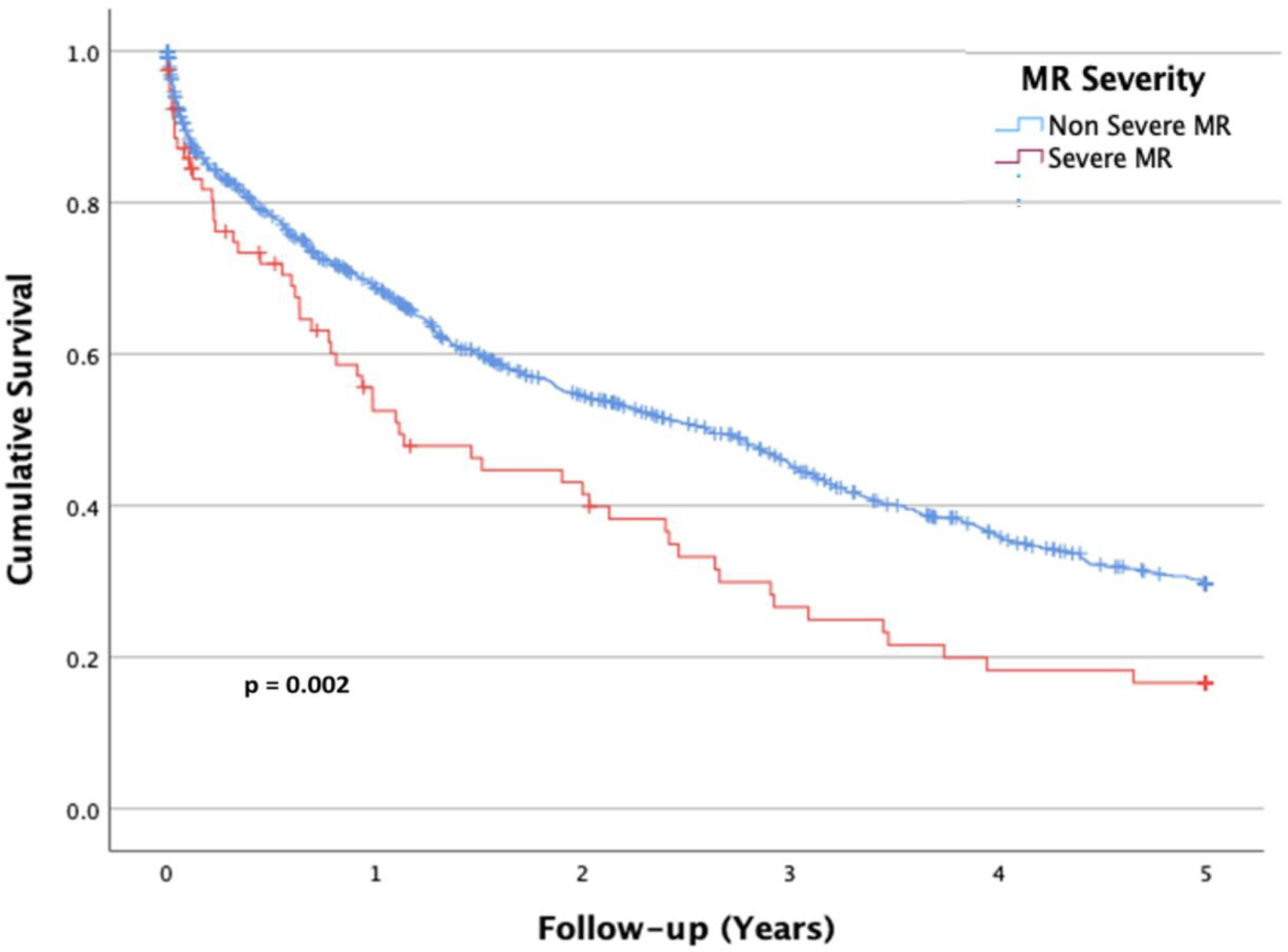
All-cause Mortality in Paradoxical Low-flow, Low-gradient Severe Aortic Stenosis by the Presence of Severe Mitral Regurgitation: Patients with severe MR had higher cumulative incidence of all-cause death than patients with non-severe MR. *MR= mitral regurgitation*.

**Figure 3:**
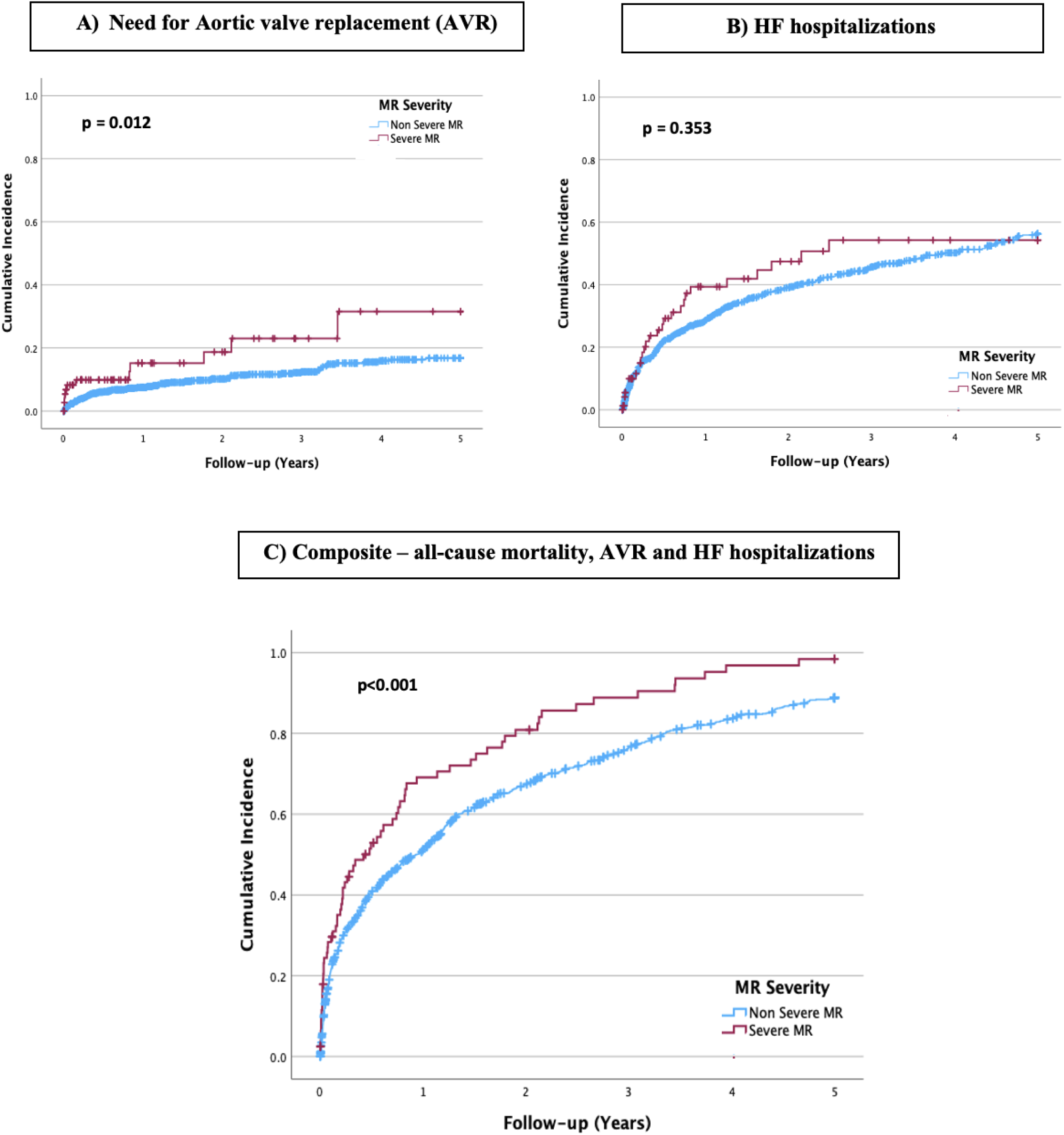
Clinical Outcomes in Paradoxical Low-flow, Low-gradient Severe Aortic Stenosis by the Presence of Severe Mitral Regurgitation: Cumulative incidences of (A) need for AVR, (B) HF hospitalizations and (C) composite outcome of all-cause mortality, AVR and HF hospitalizations in patients with paradoxical low-flow, low-gradient severe aortic stenosis, categorized by the presence of severe MR. *AVR = aortic valve replacement; HF = heart failure, MR = mitral regurgitation;*

**Table 2:**
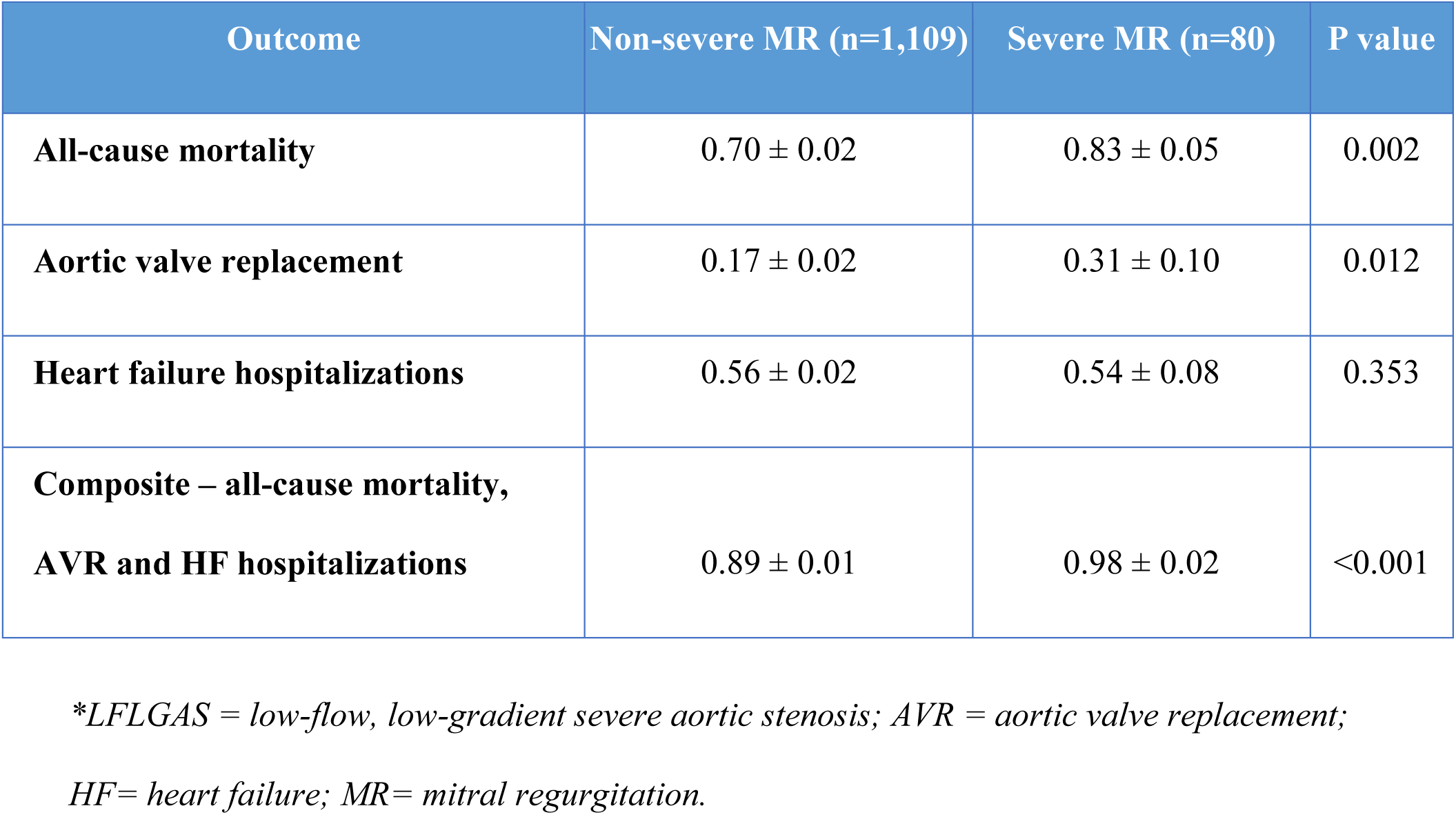
Five-year Cumulative Incidences of Clinical Outcomes in Paradoxical LFLGAS with and without Severe Mitral Regurgitation.

| Outcome | Non-severe MR (n=1,109) | Severe MR (n=80) | P value |
| --- | --- | --- | --- |
| All-cause mortality | $0.70 \pm 0.02$ | $0.83 \pm 0.05$ | 0.002 |
| Aortic valve replacement | $0.17 \pm 0.02$ | $0.31 \pm 0.10$ | 0.012 |
| Heart failure hospitalizations | $0.56 \pm 0.02$ | $0.54 \pm 0.08$ | 0.353 |
| Composite – all-cause mortality, AVR and HF hospitalizations | $0.89 \pm 0.01$ | $0.98 \pm 0.02$ | <0.001 |

*\*LFLGAS = low-flow, low-gradient severe aortic stenosis; AVR = aortic valve replacement;*
*HF= heart failure; MR= mitral regurgitation.*

Accordingly, severe MR was associated with an increased risk of mortality (HR=1.53, 95% CI: 1.16-2.01, p=0.002), AVR (HR=2.11, 95% CI: 1.16-3.83, p=0.015), and the composite outcome (HR=1.64, 95%CI: 1.27-2.12, p<0.001), compared to patients with non-severe MR. No significant difference between the groups was observed in the risk of HF hospitalizations (HR=1.21, 95%CI: 0.82-1.76, p=0.354).

The results of multivariate analysis demonstrated that patients with severe MR had a ∼1.5-fold higher risk of mortality (Adjusted HR [AdjHR]=1.43, 95% CI: 1.08-1.89, p=0.011), compared to patients with non-severe MR (Table 3). Additionally, after adjusting for clinical characteristics and the propensity score for AVR, this group exhibited an increased risk of composite outcome (AdjHR=1.64, 95% CI: 1.27-2.12, p<0.001) (Table 4). Unique predictors of adverse outcomes in the Cox proportional hazards model were older age, male sex, diabetes mellitus, history of stroke or heart failure and moderate or greater tricuspid regurgitation.

**Table 3:** All-cause Mortality Risk by the presence of Severe MR.

|  | HR | (95% CI) | p |
| --- | --- | --- | --- |
| **Severe MR vs non-severe MR** | **1.43** | **(1.08-1.89)** | **0.011** |
| Sex, Male | 1.22 | (1.05-1.43) | 0.011 |
| Age, one-year increase | 1.39 | (1.17-1.64) | <0.001 |
| Hypertension | 0.93 | (0.75-1.15) | 0.514 |
| Diabetes Mellitus | 1.28 | (1.09-1.49) | 0.002 |
| History of Stroke | 1.54 | (1.21-1.98) | <0.001 |
| History of HF | 1.98 | (1.65-2.38) | <0.001 |
| Moderate or greater TR | 1.47 | (1.23-1.75) | <0.001 |
| Hypercholesterolemia | 0.79 | (0.67-0.92) | 0.003 |

\*Cox proportional hazard models for time to all-cause death, adjusted for clinically relevant variables. *MR = mitral regurgitation; HF = heart failure; TR = tricuspid regurgitatio; AVR= aortic valve replacement;*

**Table 4:** Composite Cardiovascular Outcome Risk by the Presence of Severe MR.

|  | HR | (95% CI) | p |
| --- | --- | --- | --- |
| **Severe MR vs w/o severe MR** | **1.64** | **(1.27-2.12)** | **<0.001** |
| Predicted probability (propensity score) | 1.75 | (0.90-3.39) | 0.097 |
| Sex, Male | 1.15 | (1.01-1.33) | 0.048 |
| Age, one-year increase | 1.12 | (0.93-1.35) | 0.215 |
| Hypertension | 1.01 | (0.83-1.22) | 0.92 |
| Diabetes Mellitus | 1.23 | (1.06-1.43) | 0.005 |
| History of Stroke | 1.37 | (1.09-1.71) | 0.007 |
| History of HF | 1.99 | (1.69-2.33) | <0.001 |
| Moderate or greater TR | 1.26 | (1.09-1.47) | 0.002 |
| Hypercholesterolemia | 0.99 | (0.86-1.15) | 0.926 |

\*Cox proportional hazard models for the composite outcome of all-cause mortality, AVR and HF hospitalizations, adjusted for clinically relevant variables. *MR* = mitral regurgitation; *HF* = heart failure; *TR* = tricuspid regurgitation; *AVR*= aortic valve replacement;

### Impact of AVR on outcomes

The impact of AVR (SAVR or TAVR) on outcomes was examined. For this analysis, patients were further categorized based on whether they underwent AVR (n=261) or did not (n=928) (Supplemental Table 1). Of those who underwent AVR, 132 patients had SAVR, 124 patients had TAVR, and 52 had surgical mitral valve repair/replacement (MVR) (51 of these undergoing simultaneous SAVR+MVR). Additionally, 5 patients had transcatheter edge-to-edge repair (TEER), 1 had a transcatheter mitral valve replacement (TMVR), and 1 patient underwent simultaneous TAVR+TEER. The 51 patients who underwent mitral valve interventions (either mitral valve surgery or transcatheter mitral valve procedures) were either excluded or censored, based on the timing of the mitral valve intervention relative to the AVR. Specifically, those who had the mitral valve intervention before the AVR were excluded, while those who had it after the AVR were censored.

Data on MR mechanism was available in 191/261 patients: 75 patients (39.3%) had primary MR and 22 patients (11.5%) had secondary MR. In 46 patients (24%) the mechanism was mixed, and in 48 patients (25.1%), the MR was mild/physiologic (Table 5).

**Table 5:**
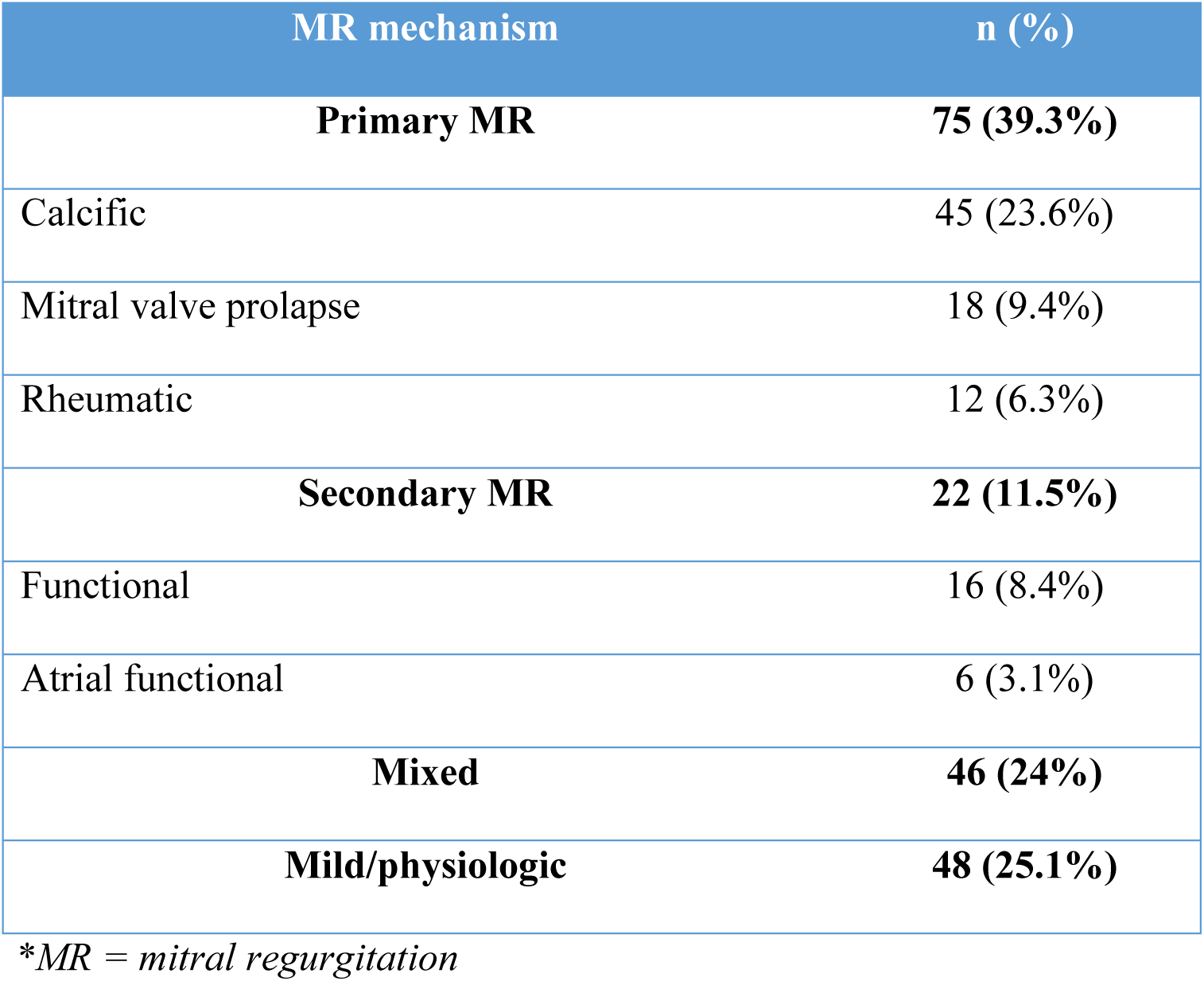
Mitral Regurgitation Mechanisms in Patients with Paradoxical LFLGAS Undergoing Aortic Valve Replacement.

AVR was associated with a reduction in mortality rates (39.8% vs 68.9%) and the overall cumulative mortality (0.50 vs 0.82, p<0.001) in the entire cohort (Figure 4a). Importantly, AVR improved survival across all grades of MR, even mild (43.0% vs 68.4%, p<0.001), moderate (38.2% vs 67.8%, p<0.001), and severe (35.3% vs 79.4%, p<0.001) (Figure 4b-d). The results of the univariate analysis showed that AVR resulted in a reduced mortality risk in the entire cohort (HR=0.39, 95% CI: 0.31-0.47, p<0.001). Additionally, AVR was clearly associated with a reduction in mortality risk in each of the MR severity groups: mild (HR=0.47, 95% CI: 0.34-0.65, p<0.001), moderate (HR=0.36, 95% CI: 0.27-0.47, p <0.001), and severe MR (HR=0.26, 95% CI: 0.11-0.62, p=0.002).

**Figure 4:**
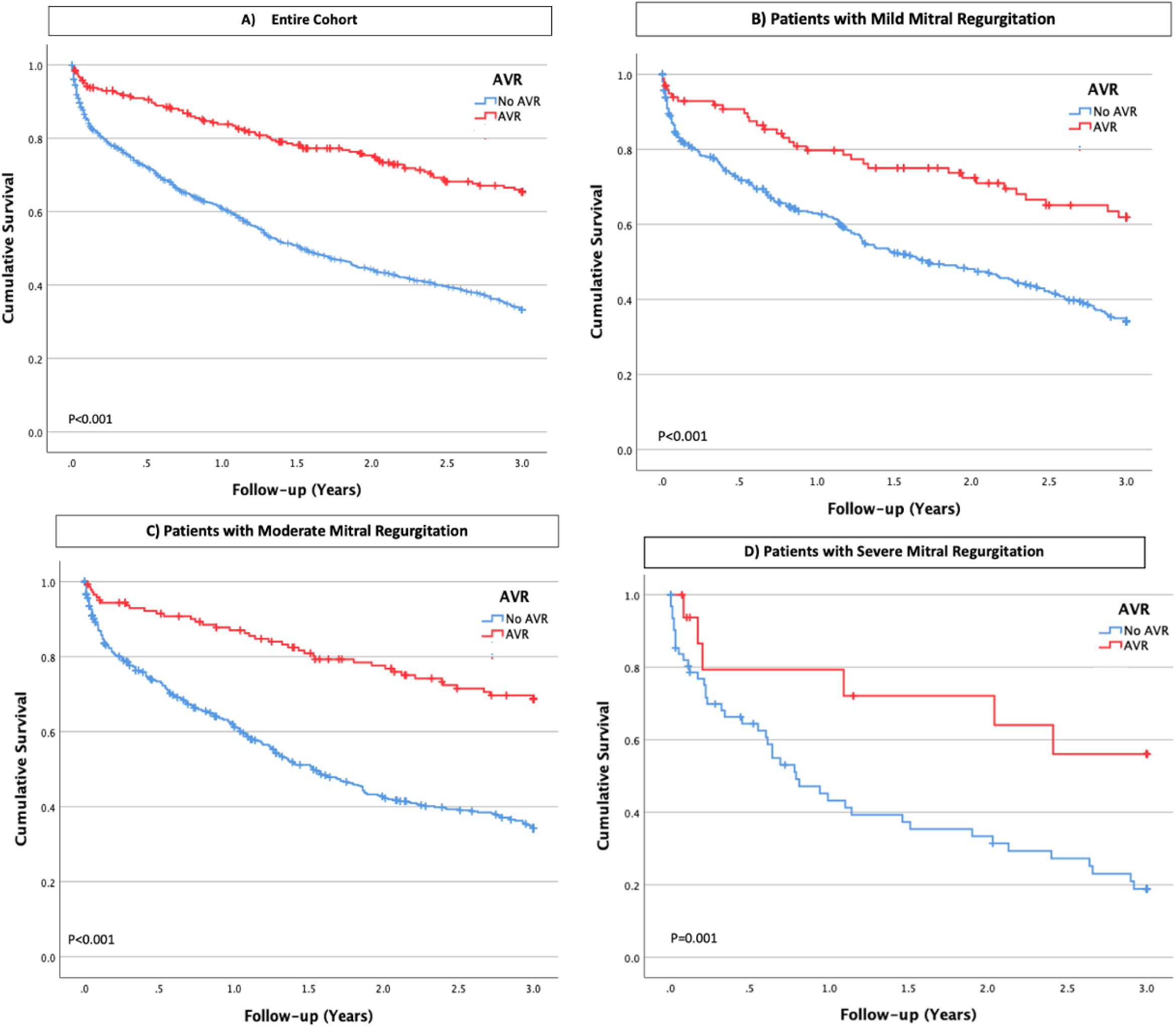
Aortic Valve Replacement Impact on Survival in Paradoxical LFLGAS: The association between surgical/transcatheter AVR and overall survival in (A) the entire cohort, and patients with (B) mild, (C) moderate, and (D) severe MR in paradoxical low-flow, low-gradient severe aortic stenosis. AVR was associated with improved survival across all MR severity subgroups. *AVR = aortic valve replacement; MR = mitral regurgitation*.

This impact of AVR on survival, across all degrees of MR, was confirmed when using a multivariate analysis, for patients with mild MR (AdjHR=0.52, 95% CI: 0.37-0.73, p<0.001), moderate MR (AdjHR=0.39, 95% CI: 0.29-0.54, p<0.001), and with the most substantial impact observed in patients with severe MR (AdjHR=0.18, 95% CI: 0.06-0.57, p=0.004). Additionally, the results of a propensity adjusted interaction model demonstrated a stronger beneficial impact of AVR on survival with an increase in MR severity (p for interaction=0.044) (Figure 5).

**Figure 5:**
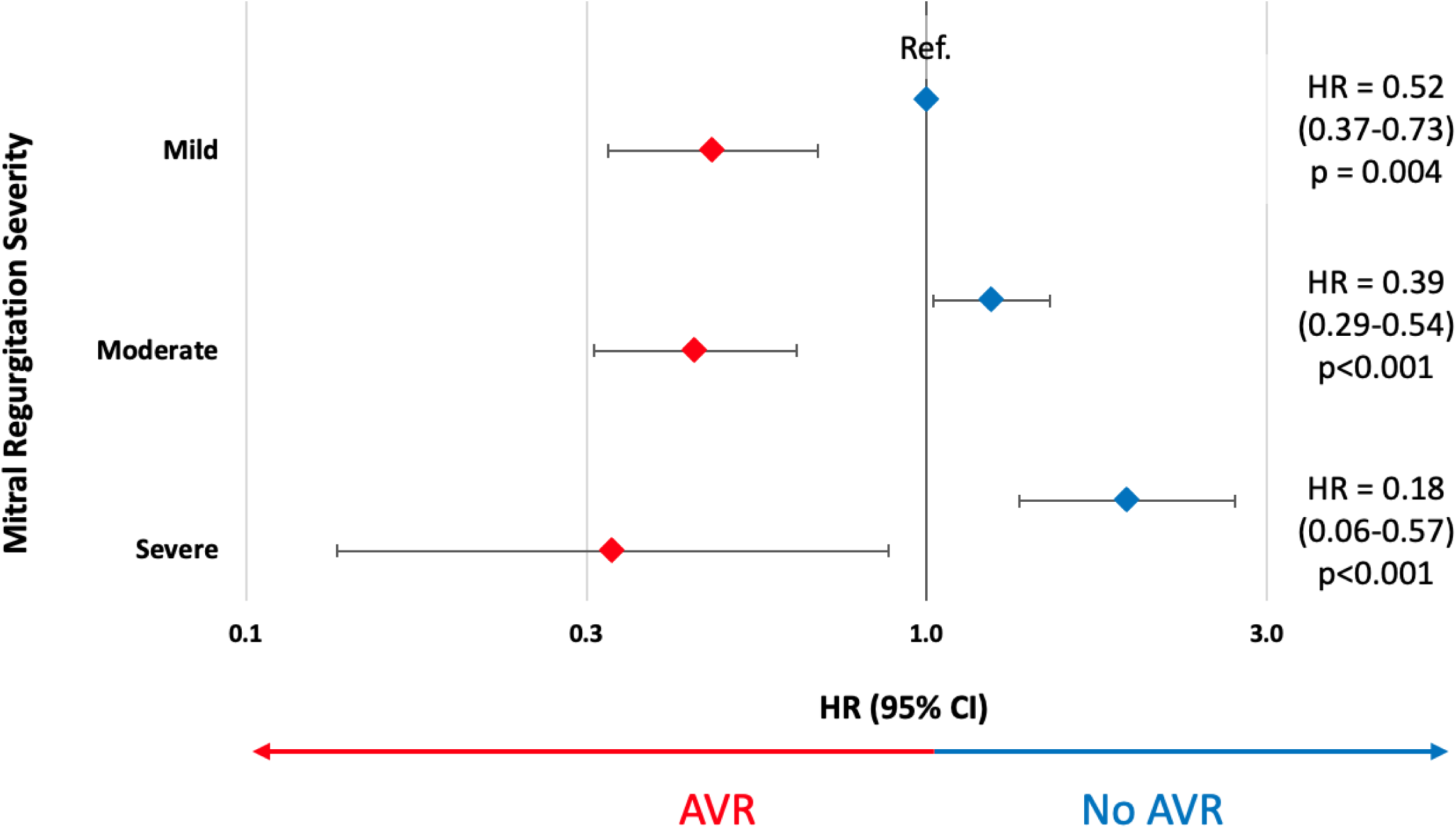
Survival Benefit of AVR by MR Severity: Results from a multivariable propensity-adjusted interaction model, demonstrating the impact of AVR on survival relative to the severity of MR in patients with paradoxical low-flow, low-gradient severe aortic stenosis. The beneficial effect of AVR on survival increases with MR severity (p for interaction = 0.04). *HR = hazard ratio; AVR = aortic valve replacement; CI = confidence interval; MR = mitral regurgitation; Ref= reference group;*

## Discussion

This study shows that patients with paradoxical LFLGAS with concomitant severe MR have a significantly increased risk of all-cause mortality and a composite cardiovascular outcome.

Furthermore, undergoing AVR in patients with paradoxical LFLGAS was associated with improved survival across all MR severity levels, particularly with severe MR (Central Illustration). These findings provide important outcomes data in the management of patients with paradoxical LFLGAS and MR.

### Physiological implications

Low-flow, low-gradient aortic stenosis was first described in 2007 by Hachicha *et al* (1), and is a common diagnostic and therapeutic dilemma encountered in clinical practice, with a prevalence of 8-25% of patients with aortic stenosis (6,18,19). LFLGAS is diagnosed by Doppler echocardiography after carefully excluding measurements errors, and is characterized by AVA≤1 cm^2^, a mean gradient ≤40mmHg and a low flow state. In patients with preserved LVEF, several factors have been suggested as potential drivers of the low flow state. A common physiologic scenario is the presence of a small, poorly compliant LV cavity due to concentric LV hypertrophy, associated with increased ventricular afterload, and subclinical myocardial dysfunction (7,20–22). The superimposition of severe MR on this already complex physiology further decreases forward stroke volume, and has been previously shown to be an independent determinant of the low flow pattern (9,10).

#### Severe MR as a flow diverting lesion

Low flow is currently defined in the guidelines by stroke volume index ≤35 mL/m^2^ (23). However, from a physiological standpoint, low flow is more accurately defined by transvalvular flow rate (Q), which is the ratio of stroke volume to ejection time. Considering that transvalvular flow rate is influenced not only by stroke volume but also by LV ejection time, low Q better reflects the low flow state than stroke volume. Our group has previously demonstrated that flow rate is a significant factor in determining the prognostic value of AVA (13). In this current study, we defined low flow using Q rather than LV stroke volume. This approach allowed us to examine a truly paradoxical LFLGAS population, capturing the distinctive physiology of this group.

Our results show that patients with paradoxical LFLGAS and severe MR have lower flow rates compared to patients with non-severe MR. Furthermore, our data revealed a progressive reduction in flow rate with increasing MR severity. This observation emphasizes the dose-response effect of MR as a flow-diverting lesion that incrementally redirects flow away from the aortic valve.

#### Severe MR superimposed on paradoxical LFLGAS leads to differential remodeling

Patients with paradoxical LFLGAS and severe MR showed a more eccentric LV hypertrophy pattern, characterized by larger LV internal diameters, increased LV mass index and lower relative wall thickness, compared to patients with non-severe MR. This eccentric type of hypertrophy can be viewed as a compensatory mechanism to the volume overload caused by the MR, especially when superimposed on top of the small LV characteristic of patients with LFLGAS, that is already lacking preload reserve.

### Clinical implications

#### Severe MR in paradoxical LFLGS is associated with worse clinical outcomes

The unique pattern of paradoxical LFLGAS with concomitant severe MR not only sheds light on the inconsistencies in aortic stenosis severity classifications but is also potentially associated with adverse outcomes. However, large clinical trials evaluating outcomes in paradoxical LFLGAS have excluded patients with moderate or severe MR (1,3,4,24–26). This exclusion stems from the complexity introduced by simultaneous significant valvular abnormalities. Yet in clinical practice, mixed valve disease is fairly common and warrants more frequent and thorough investigation.

In our study, patients with paradoxical LFLGAS and severe MR exhibited reduced overall survival and a worse composite cardiovascular outcome, compared to patients with non-severe MR. The higher LV mass index in these patients suggests that the combination of severe pressure and volume overload, induced by paradoxical LFLGAS and MR, respectively, leads to excessive cardiac hypertrophy. This hypertrophic response may contribute to the mechanisms explaining the lower survival observed in these patients.

Earlier observational studies exploring the prognostic significance of MR in patients with classical LFLGAS (defined by LVEF<50%) did not consistently identify an association between baseline moderate or severe MR and adverse outcomes in those undergoing TAVR, as evidenced by the multicenter TOPAS-TAVI registry (27). A recent study by Ferruzi *et al* encompassing both classical and paradoxical forms of LFLGAS, demonstrated that moderate to severe MR in individuals undergoing TAVR predicts an unfavorable outcome within the first year (28).

Notably, our study uniquely assesses the impact of severe MR on outcomes in a pure population of paradoxical LFLGAS, a population not previously explored.

#### Aortic valve replacement is associated with improved survival in paradoxical LFLGAS, especially among patients with concomitant severe MR

The association between severe MR and increased mortality risk in paradoxical LFLGAS makes physiologic sense, as MR exacerbates the low flow state. However, what is noteworthy and deserves emphasis, is the novel finding that there appears to be a survival benefit with AVR, which increases with the severity of MR. It has been previously shown that the prognosis of paradoxical LFLGAS patients improves with AVR (23). Consistent with this, our study showed that AVR was associated with improved survival in the entire cohort, and this beneficial effect was present across all degrees of MR. Interestingly, patients with severe MR experienced better outcomes following AVR than those with lower degrees of MR. This was observed even after adjusting for numerous confounding factors and excluding those who underwent mitral valve interventions. These findings suggest that AVR may alter the natural progression of paradoxical LFLGAS with severe MR, and may be beneficial earlier in the disease course.

In the current landscape of increasing transcatheter options for valvular heart disease, our results potentially contribute to an evolving paradigm of a more tailored approach to patients with multi-valvular disease. Unlike the historical practice of addressing all valves diseases simultaneously during open-heart surgery, today’s interventions enable a staged approach individualized to patient’s symptoms and valve disease. This allows for a nuanced understanding of how each intervention impacts outcomes, offering us the possibility to separate and specifically address how AVR influenced outcomes in this distinct patient population.

#### Study limitations

Inherent limitations of this study arise from its retrospective design. However, the institutional echo database is based on regular quality assessments to ensure accuracy and reproducibility. Our database offers the benefit of enabling extensive data evaluation and the analysis of real-world, all-comers participant data. Also, to the best of our knowledge, this is the first study to report the effect of severe MR and AVR on clinical outcomes among patients with paradoxical LFLGAS (with a rigorous definition of low flow state).

We did not have information on symptoms or the New York Heart Association (NYHA) classification at the time of index echocardiography, which limited our ability to link symptoms onset to patient outcomes. Nonetheless, we assumed that most patients with coexisting two significant valve disorders will exhibit some degree of symptoms, making it challenging to discern the specific valve disease responsible for these symptoms. Lastly, we did not incorporate aortic valve leaflet calcium score data and STS scores, but these were considered during the heart team evaluation of the patients included in our study. While calcium score data could offer additional anatomical insights for assessing aortic stenosis severity and guide procedural decision making, our analysis relied on echocardiographic parameters and hemodynamic measures diagnosis.

## Conclusions

In conclusion, this study contributes to our understanding of paradoxical low-flow, low-gradient severe aortic stenosis and severe MR. Our data demonstrate that the coexistence of these two conditions leads to reduced overall survival and worse composite cardiovascular outcomes.

Importantly, we found that aortic valve replacement was associated with improved survival rates in patients with all grades of MR, especially among those with severe MR.

## Data Availability

All data is available through the corresponding author.

## Non-standard Abbreviations and Acronyms

AVA: aortic valve area
AVR: aortic valve replacement
HF: Heart failure
LFLGAS: low-flow, low-gradient severe aortic stenosis
LV: left ventricle
LVEF: left ventricular ejection fraction
MR: mitral regurgitation
SAVR: surgical aortic valve replacement
TAVR: transcatheter aortic valve replacement

## Central Illustration: Mitral Regurgitation in Paradoxical LFLGAS

In patients with paradoxical LFLGAS, MR acts as a flow-diverting lesion, decreasing forward flow across the aortic valve and leading to worse clinical outcomes. Undergoing AVR is associated with improved survival across all degrees of MR, particularly in patients with severe MR. *AVR = aortic valve replacement; LFLGAS = low-flow, low-gradient severe aortic stenosis; MR = mitral regurgitation*.

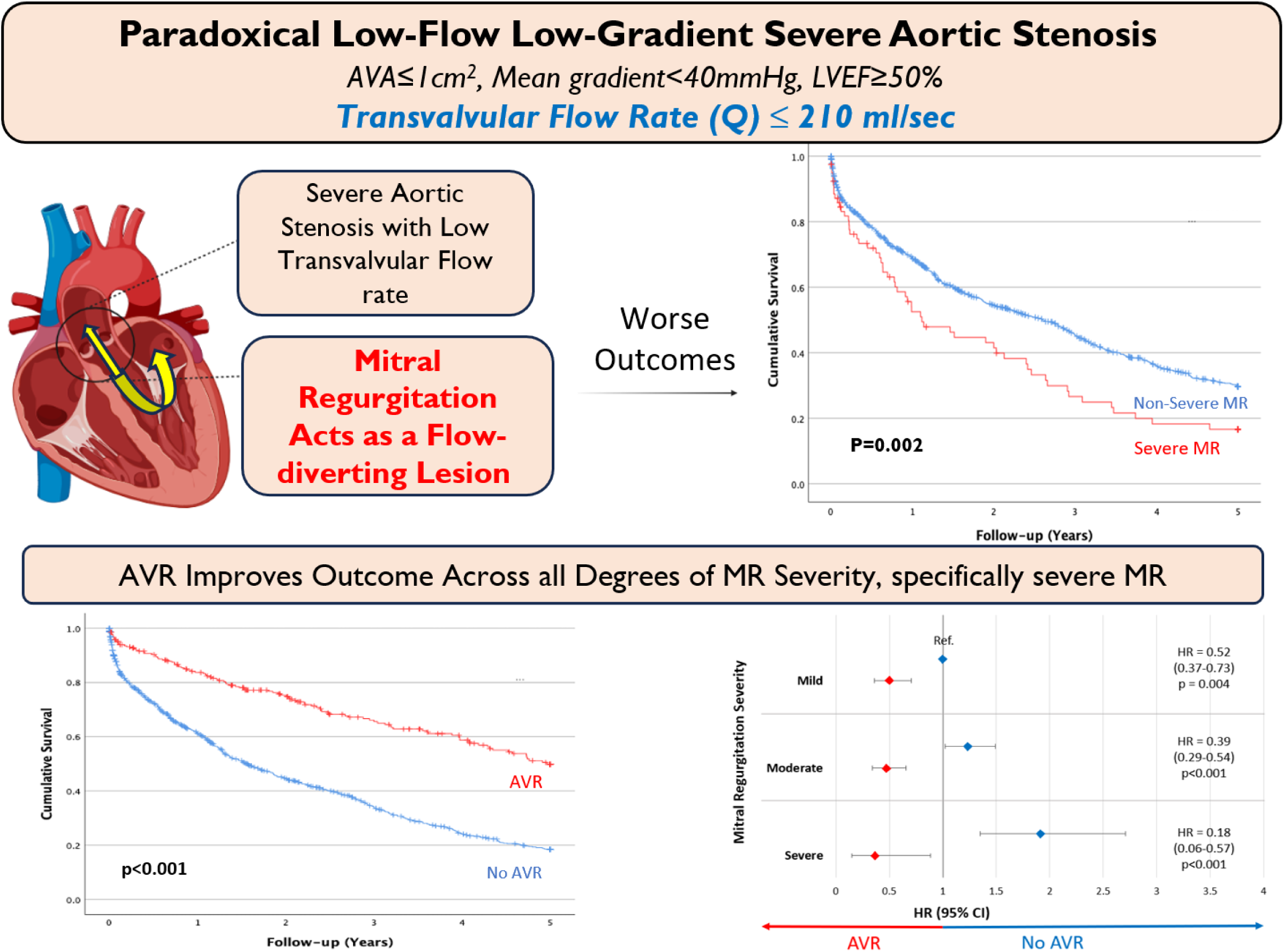

